# Qualitative protocol to support detection of the early presentation and diagnosis of mycetoma in Sudan

**DOI:** 10.1101/2023.08.17.23294207

**Authors:** Caroline Ackley, Victoria Hall, Eiman Siddig Ahmed, Natalia Hounsome, Mohamed Nasr Elsheikh, Shahaduz Zaman, Sahar Bakhiet

## Abstract

The neglected tropical disease (NTD) mycetoma is a chronic and progressively destructive infectious disease endemic in Sudan. There is a growing body of scientific research on mycetoma -causes, transmission, treatment, and impact from a clinical and biomedical perspective. However, there is further need for in-depth medical anthropology research on the disease to successfully translate biomedical advances into elimination and control programmes. Given this background the NIHR Global Health Research Unit on NTDs at Brighton and Sussex Medical School is leading multidisciplinary research on skin NTDs, including mycetoma, with a medical anthropology research component exploring how community engagement can lead to earlier presentation of mycetoma in Sudan. This protocol paper sets out the research aims and approaches to generate new knowledge on mycetoma in Sudan once the political situation becomes stable and it is safe to carry out global health research once again. We continue to develop appropriate community engagement intervention strategies, while activities like training and capacity strengthening get underway remotely.

## Introduction

Neglected tropical diseases (NTDs) affect more than one billion people globally, with skin NTDs affecting hundreds of millions of people (1) Of the 20 NTDs listed by the WHO, more than half present with skin manifestations (2). The WHO’s list of skin NTDs includes Buruli ulcer, cutaneous leishmaniasis, post-kala-azar dermal leishmaniasis, leprosy, lymphatic filariasis (lymphoedema and hydrocele), mycetoma, onchocerciasis, scabies, yaws, and fungal diseases (ibid.). Skin NTDs can cause discomfort, stigma, physical disabilities, disfigurement, mental distress, loss of social status, and affect the quality of life of those afflicted (3) (4).

Mycetoma is a skin NTD endemic in countries in Africa, Asia, Europe, and Latin America with the causative organisms distributed along tropical and subtropical areas called the ‘Mycetoma belt.’ Most cases have been reported in Sudan and Mexico (5), with most of Sudan’s cases predicted to be in the central and southeastern states (6). The southeastern state where this study is located, Sinnar State, reported 9% of countrywide cases being treated at the Mycetoma Research Centre (MRC), Khartoum, Sudan between 1991 and 2018 (6).

Mycetoma is a chronic and progressively destructive infectious disease thought to be acquired by traumatic inoculation of fungi or bacteria into the subcutaneous tissues that spreads to affect the skin, deep tissues, and bone (5). The organisms responsible for mycetoma are thought to originate in soil or animal dung, with both bacterial and fungal causative agents identified in soil samples (7), and fungal agents identified in dung (8). Both causative agents are introduced into subcutaneous human tissue through open wounds (8,9). It is usually affects the foot, but any part of the body can be affected. Wearing open-toed sandals or walking barefoot while attending to livestock is common in Sinnar State, and can result in frequent thorn pricks/cuts, subsequently leading to high rates of mycetoma (10). People of low socioeconomic status and those whose main source of income is agriculture or herding are the most affected (11).

To tackle the burden of mycetoma the WHO calls for active case finding with early diagnosis and treatment, along with further public health actions, including ‘reinforcing awareness among affected communities’ (2). This was part of the WHOs return to the declaration of Alma-Ata of 1978 (12) which emphasized community involvement, volunteer activities, relationships between local governments and communities, and decentralization (13). Our proposed research responds directly to this by exploring if community engagement can lead to earlier presentation of mycetoma in Sudan, and more specifically in Sinnar State.

### The need for social science and community engagement in the study of mycetoma

Global health research incorporates epidemiological perspectives along with the social sciences, and it recognises that many different types of research contribute to policy, practice, and future research (14). Biomedical technologies are critical to managing NTDs, however, without efforts to also address the social, cultural, economic, political and environmental factors that affect populations people will continue to live in neglect and poverty (15). As such, there is a strong need for social science research on mycetoma and other NTDS. More specifically, a review by Manderson, et al (13) into the ‘current status of applied social science research in tropical disease control’ identified several key research themes, including community participation in control programs and the need for a shared commitment between people and their government. In what follows we situate this study in the NIHR Research Unit on NTDs and alongside social science research on mycetoma.

#### NIHR Research Unit on NTDs

This study is part of the program of research conducted by the NIHR Research Unit on NTDs at Brighton and Sussex Medical School (BSMS). Phase I of the Unit was conducted from 2018-2021. Below we detail key findings from Phase I that have informed our more recent work in Phase II (2021-26). It also sits alongside the Social Sciences for Severe Stigmatising Skin Conditions (5-S) Foundation also at BSMS (16). The 5-S foundation examines the cultural, social, economic, and policy contexts of podoconiosis, mycetoma, and scabies in relation to the dynamics and dimensions of stigma. This study differs from the aims of the 5S Foundation in that it specifically focuses on the role community engagement can have in the early presentation and diagnosis of mycetoma in Sudan.

##### Phase I

During Phase I of the NIHR project we conducted anthropological research aimed at understanding perceptions and experiences of mycetoma in Sudan; namely those of mycetoma patients who attend the Mycetoma Research Centre (MRC), and within the Wad Elnimear community in Sinnar State. Anthropological research was conducted in Wad Elnimear and the MRC, and focus group discussions were held in Sinnar (17). The workshops were meant to inform an intervention strategy; however, the study timeline was delayed due to COVID-19 and a military coup in Sudan. We also carried out several capacity strengthening activities, which further reinforced our research network in Sudan.

###### Anthropological research

A number of independent research projects, many conducted by the MRC, have contributed to our team’s overall body of data during Phase I of our group research (6,9,11,18–21); this has included a PhD project on mycetoma in endemic regions of Sudan, and their current intervention strategies and treatment gaps (17). This thesis concentrated on socio-cultural and political-economic dimensions of mycetoma in Sudan, the research was predominantly qualitative, and included aspects of formal and informal interviewing; observation and conversational interaction; and periods of more “traditional” ethnographic research. Elsheikh was specifically interested in locating possibilities for new forms of intervention - he wished to draw attention to the opportunities available for more co-creative forms of intervention methodology and strategy in rural Sudan. This has also been a key focal point for the research of the mycetoma research team more generally, and, as such, this emphasis on the dynamics of intervention has informed much of our research during Phase I - in addition to prompting many new questions to be addressed during Phase II.

In particular, Elsheikh’s doctoral research has contributed to our foundational knowledge of mycetoma in Sudan. He identified treatment pathways, cultural concepts (10), and beliefs in mycetoma management. Specifically, his findings indicate that traditional healers are crucial players in the detection and management of mycetoma, as they are often the first point of contact for most mycetoma patients and often a source of delay in disease identification and, sometimes, the only choice for patients when treatment is expensive. This is an area of particular interest and importance that we feel will benefit substantially from further research during Phase II of this project.

In close alignment with his research on traditional healers and their methods, Elsheikh also argued for a new understanding of the dynamics of stigma surrounding mycetoma and how these may impact on the capacity of health-seeking behaviours for individuals afflicted by mycetoma. Elsheikh mentioned the importance of stigma throughout his research. He also detailed how stigma is a variable - and often unpredictable - phenomenon. It can affect not only the likelihood of people being able to seek-out care from traditional healers, but it can also be influenced (either positively OR negatively) by accepting such treatments when they are made available or are offered by healers. Depending on circumstance and/or context, stigma can be alleviated by treatment from traditional healers, or it can, equally, be induced or exacerbated. This requires further ethnographic research during Phase II of this project.

###### The current situation in Sudan and how this has impacted our research

Recently, Sudan has struggled with a complex and challenging political, social, and economic situation. The country has experienced significant political upheaval. Since 2018, planned protests resulted in a revolution, which ultimately led Sudan’s ex-president Omar al-Bashir to be overthrown. Following that, a transitional government was established to pave the way for democratic elections and address long-standing governance issues, economic disparity, and social justice (22). After three years, and on October 25, 2021, the Sudanese Armed Forces (SAF) deposed the transitional government, and Abdel Fattah al-Burhan, the Sudan army chief, took over. He was seconded by Mohammed Hamdan ‘Hemedti’ Dagalo as his deputy in the newly formed government. Hemedti also happens to be the from the paramilitary Rapid Support Forces (RSF) (23).

Then, in April 2023, a sudden act of violence and war erupted in Sudan between the Rapid Support Forces (RSF) and the Sudanese Armed Forces (SAF) because of a disagreement between the two forces in different parts of Sudan, particularly in Khartoum and Darfur. According to available information, as of June 20, 2023, the conflict has resulted in an estimated death toll of 3,000 to 5,000 individuals, with an additional 6,000 to 8,000 sustaining injuries. Moreover, by July 2023, approximately 2.2 million individuals have been displaced within Sudan, while 645,000 others have sought refuge in neighboring countries (23).

The turmoil of the last decade has greatly impacted the healthcare system in Sudan (24). The healthcare system has suffered from many challenges including and not limited to a leadership vacuum, suspended service delivery, diminished human resources for health, disrupted medical supply chain, and interrupted health funding. This is all in addition to the burden of the COVID-19 pandemic in conjunction with continuous mass protests, pushing the healthcare system to the verge of collapse (25).

Despite the more recent ceasefire agreements, violent bombing has continued thus affecting civilians, alongside an intentional attacking of hospitals and healthcare personnel through random shootings, missiles and artillery (26). In addition, 70% of the hospitals in Khartoum and other states are closed and not functioning due to the continuous attacks against health staff and those sheltering in health facilities (27).

Due to this serious collapse in the health system and the lack of security, many projects have paused their work in Sudan; including the NIHR Global Health Research Unit (GHRU) on NTDs at BSMS. The NIHR continues to fund Sudanese researchers, including PhD students, while projects are redesigned or paused. The NIHR GHRU intends to resume as soon as the situation stabilizes in Sudan, and regional peace and security is ensured. The GHRU intends to conduct a comprehensive assessment of the health and social challenges faced during this period of instability. It will also facilitate the development of targeted interventions for people with mycetoma in Sudan. We hope that future research findings will not only contribute to the disease knowledge-base, but also play a vital role in shaping policies and strategies aimed at improving the overall well-being of the Sudanese population in the aftermath of instability.

When it becomes safe to continue research, we have identified three themes that need further exploration in order to develop a multifaceted intervention that targets the early presentation and diagnosis of mycetoma: (1) further understanding the disease and those most affected, (2) traditional healers and biomedicine, and (3) stigma.

1) <u>Further understanding the disease and those most affected by it</u>: In Phase I, we observed different health seeking pathways for people affected by mycetoma, which often led to delayed presentation of the disease when people finally sought medical attention at the MRC. Additionally, some patients sought support from traditional healers even after receiving treatment at the MRC. We now seek to explore the knowledge, attitudes, and perceptions (KAP) of mycetoma amongst those living in mycetoma endemic areas. Principally, we will be asking: *What is people’s knowledge of mycetoma and how it is contracted? What is their attitude towards preventative measures/an intervention? And, how is the risk of the disease perceived?* We therefore also s eek to explore “knowledge awareness”, as well as the more generalised levels of knowledge *and* awareness of those in endemic areas. “Knowledge awareness” refers to an individual’s state of being informed and having perceived information about others’ knowledge. This leads us to *then* ask: *who is perceived as having knowledge on mycetoma, and who is able to treat mycetoma?*

During Phase II, we will identify which populations have a gap, or gaps, in their knowledge of mycetoma, and who might, therefore, be best targeted for preventative measures/an intervention. To do this, we will conduct varied stakeholder mapping to learn who the community leaders are, and who might serve as gatekeepers and facilitators for an intervention. We will also attempt to gain insight into who are the most marginalised members of these societies (including women and children). This is to ensure that any of our planned intervention activities include those most in need, or formerly overlooked populations. Finally, we will conduct asset mapping to identify the resources and skills of individuals, groups, and organisations in the study site(s). Asset and stakeholder mapping will allow us to develop an intervention that is sustainable and ‘comes from within;’ meaning it uses local knowledge, skills, and resources and can be led by community leaders. We hope this approach facilitates an intervention that ‘lives’ beyond the project timeline through local capacity.

2) <u>Traditional healers and biomedicine</u>: In Phase I we learned that there are several parallel paths of health-seeking behaviour(s), often with people going to the traditional healers or religious healers first and to their (biomedically trained) doctor, or to the MRC, second. Seeking biomedical care as a second option results in a late presentation of mycetoma, and, as such, to much more adverse health outcomes (see, e.g., Elsheikh 2022). Traditional healers recite the Quran, use prayer, and/or utilise varied herbal medicines to treat mycetoma. Religious healers are sheikhs (traditional healers can also be sheikhs, and sheikhs are considered to be community leaders and thus influential) who use prayer or ‘*dua*’ to heal. They sometimes organise *Alholia*, or large prayer gatherings, for healing various ailments and problems. Praying in large groups means that one’s prayer or specific request for help is more likely to be heard. Our foundational research on traditional and religious healing at times conflates the two, so we propose further exploration to understand the role religious healing and prayer plays in health seeking behaviours and late presentation of mycetoma.

Some people prefer to use one system of health management at a time, while others visit the traditional healer (and/or religious healer) and biomedical doctor simultaneously. It is common for people to visit the biomedical doctor after having been referred by their traditional healer or if traditional medicine fails to cure them; however, there are many practical reasons that people prefer to get help from their traditional healer first, including: cost effectiveness, distance^1^, and frustration with duration of treatment and treatment outcome from biomedical doctors. Few people seek traditional healers after being treated by biomedical doctors. Ultimately, we want to understand health-seeking pathways, and the relationships between all individuals involved in these pathways (regardless of role or perceived societal “level”), in order to facilitate productive conversations between stakeholders, and to encourage earlier biomedical health-seeking behaviours amongst affected populations.

3) <u>Stigma</u>: In Phase I, and through the ongoing research of our colleagues in the 5-S Foundation, we have learned that people with mycetoma experience stigma, and yet they are still incorporated into the social fabrics of their community(ies). Bakhiet et al. (2018) found that mycetoma related stigma is most felt amongst women and children. Additionally, Fahal et al. (2014), Bakhiet et al. (2018), Zijlstra et al. (2016) and Abbas et al. (2018), have *all* indicated that women and children are less likely to receive medical care due to stigma, as well as financial, and gender-role considerations. This is despite men and women being at similar risk of exposure (28). Late presentation often means a more progressive form of the disease and increased need for surgical involvement. This can then lead to lifelong disability, and to further burdens and subsequent stigma on those diagnosed and their families. This also leads to higher rates of school dropout for afflicted children/young adults, negative impacts on marital status and employment, and to individual and/or familial vulnerability to heightened psychological distress. Elsheikh (2022) found that it is believed that stigma is a genetic disease, transmitted from within, meaning that not only is mycetoma believed to be genetic (although it is an infectious disease, cases can cluster in families leading to this belief), but that stigma and vulnerability can also be inherited or “passed on” (29). This stigma affects individuals suffering with mycetoma, as well as entire communities labelled as “mycetoma villages”. We seek to gain further understanding into the experiences of felt, enacted, and structural stigma in endemic areas, and of how such stigma then often leads to late presentation. We will also explore the belief that stigma itself is a genetic disease. We want individuals and communities to have the opportunity to share their experiences of stigma, and for others to learn about the impact stigma has on affected individuals. We will use our findings to inform healthcare providers and researchers, providing qualitative and quantitative research data and other evidence as available, so as to then reduce *their* impact on structural stigma. This is an important “layer” for successful intervention development and implementation.

###### Health economics

Our recent cross-sectional population study conducted in Sinnar State, Sudan (21) showed that economic factors such as agricultural practices and reduced access to sanitation and clean water contribute to the development of mycetoma. Poor access to health care, long travel distances and substantial financial costs were barriers to seeking treatment for mycetoma.

In this study, we will focus on assessing health-related quality of life and the use of healthcare services by people with mycetoma. Addressing these two components is essential for the cost-effectiveness analysis of future interventions to improve detection of mycetoma.

## Principal Research question

Can community engagement support detection of the early presentation and diagnosis of mycetoma in Sudan?

This is a complex, highly-layered, and often both circumstantial and context-dependent question. As such, we have identified several equally crucial sub-research questions, so as to draw attention to this nuance, relationality, influence, specificity, and, in some cases, temporality or unpredictable tendency to change.

As a preliminary point of departure, these sub-questions are as follows. They shall be further developed and/or altered as appropriate alongside our research and acquired data throughout Phase II.

### Sub-Research questions

1. Understanding the disease and stakeholders:

1. What is people’s knowledge of mycetoma and how it is contracted?
2. What is their attitude/attitudes towards preventative measures/an intervention?
3. How is the risk of the disease perceived?
4. Who is perceived as having knowledge on mycetoma, and who is perceived as being able to treat mycetoma?
5. How can increased insight into mycetoma KAP (above) and knowledge awareness be used to improve health communication and to lead to early presentation and diagnosis?
2. Traditional healers and biomedicine:

1. What are patients’ current health seeking pathways?

1. Is religious healing a distinct pathway? What role, if any, does it play in health-seeking behaviour and the late presentation of mycetoma?
2. What is the relationship between traditional healers and biomedical doctors in treating mycetoma?
3. What are the points of tension and convergence?
4. How can photovoice possibly facilitate conversations between traditional healers, biomedical doctors, and patients - ultimately leading to early/earlier presentation?
3. Stigma:

1. How is stigma experienced, particularly by women and children?
2. In what ways is ‘stigma a genetic disease’ (El Sheikh, 2022), and what impact does this have on individual experiences of stigma and early presentation/diagnosis, or the prevention of early presentation/diagnosis?
3. How do medical doctors and researchers contribute to structural stigma, and how can this be addressed (potentially by our intervention strategies, as well as by other methods)?
4. Does using theatre to share experiences of stigma reduce its impact and lead to early presentation and diagnosis?
4. Health economics:

1. What resources are required to deliver the intervention and what are their costs?
2. Can we measure changes in health-related quality of life in people with mycetoma using the World Health Organization tool WHOQOL-BREF?

## Materials and Methods

The three themes and their corresponding research and intervention activities will be carried out by a research team consisting of: a senior researcher, two UK-based postdoctoral research fellows, one Sudan-based postdoctoral research fellow, and one-Sudan based PhD student. The three research fellows will contribute towards each theme, with the Sudan-based postdoc focusing on theme 1 ‘understanding the disease’, the Sudan-based PhD student focusing on theme 2 ‘traditional healers and biomedicine’, and two UK-based postdocs focusing on theme 3 ‘stigma’.

The research themes will be explored using different methods, some familiar, some socio-scientific, and some creative. We aim for this methodological approach to be adaptable to other countries where mycetoma is endemic, for the anthropological study of other NTDs, and for a blueprint of intervention activities that can support early detection of mycetoma (and potentially other NTDs), thus reducing disease burden.

This qualitative protocol is currently under review with the Brighton and Sussex Medical School Research Governance and Ethics Committee (RGEC), however it has not yet received ethical approval. BSMS RGEC has advised the study team that due to the current political and safety situation in Sudan it is not possible to obtain insurance from the University of Sussex to carry out the research, and therefore they cannot grant ethical approval. Once the situation stabilises we will reapply for ethical approval and insurance coverage.

### Aim

To conduct research on the three aforementioned themes, and to then develop a multifaceted intervention; implement it; and, finally, evaluate this intervention for future efficacy and the further development/improvement of future intervention strategy(ies) targeted to improve early detection of mycetoma.

### Design

This research is a mixed-methods study with ongoing data analysis based on an iterative approach. This allows us to refine our methods for each theme based on new insights. The study will also include stakeholder involvement through the establishment of a community advisory board (CAB). The CAB will provide guidance and feedback for each theme of the medical anthropology research and proposed interventions. They will also help develop a theory of change. The CAB will meet every other month for the duration of the study and will be composed of stakeholders involved in all implementation activities. Stakeholders will be identified through a stakeholder mapping exercise at the start of the study.

### Setting of the study

The study will take place in a mycetoma endemic community in Sinnar State, Sudan. According to data collected by the Mycetoma Research Centre Sinnar, Al Gaziera, While Nile, and Khartoum States have the highest number of recorded mycetoma cases (30). In Sinnar disease prevalence is found to be as high as 0.87% (28). In Sinnar the disease predominance is slightly higher in females (28), contradicting previous studies who reported the male/female ratio as 3-4:1 (31,32). Additionally, in Sinnar it is reported that lower extremities are most affected in men, while females have more hand mycetoma (28).

Sinnar not only presents interesting epidemiological findings, but it also allows us to continue Phase II in a state where we already have gatekeeper support and approval. There is also a Mycetoma Research Centre treatment centre in Sinnar, allowing us to refer cases for biomedical investigation when appropriate.

### Sample size

In order to recruit participants, we will need gatekeeper permission to enter the community. We will get gatekeeper permissions from the Mycetoma Research Centre (MRC) based on their extensive work on mycetoma in Sinnar State. The MRC will request permission from the Sinnar state Ministry of Health and the local health offices on our behalf. Once permission is granted, we will be able to enter the community and begin recruiting participants for all research and intervention activities.

Sample size will depend on the activity and participants will be recruited based on two models of saturation (33). We take data saturation to be a complex component of research validity and integrity, and therefore define it according to the models of inductive thematic saturation and data saturation (33). Inductive thematic saturation relates to the emergence of new codes or themes during analysis. Data saturation relates to the degree to which new data repeat what was expressed in previous data during data collection. Once both models of saturation have been achieved, we will stop collecting data. We provide a range of the number of participants to allow us the flexibility to achieve both models of saturation.

### Inclusion/exclusion criteria

Inclusion criteria: consents to take part (written and verbal); over age 18; resides in the study site. Exclusion criteria: does not consent to take part; under age 18; does not reside in the study site.

### Characteristics of participants and how the sample will be selected

#### Theme 1

1. Stakeholder mapping: We will recruit 20-30 people to participate in stakeholder mapping. They will be of mixed age and gender to represent the community at large. 10-15 participants will be purposively selected based on age, gender, and status in the community. The next 15-20 participants will be selected using the snowball method. Data will be collected by the PDRFs and PhD student.
2. Asset mapping: We will recruit 20-30 people to participate in asset mapping. They will be of mixed age and gender to represent the community at large. 10-15 participants will be purposively selected based on age, gender, and status in the community. The next 15-20 participants will be selected using the snowball method. Data will be collected by the PDRFs and PhD student.
3. Cultural Domain Analysis: We will recruit 100-150 participants to participate in CDA. The final number of participants will depend on data saturation. Participants will be recruited at a community-based screening run by local health centres. Patients who attend the community-based screening at the health centre will be recruited to participate in CDA. As patients wait for the screening, they will be recruited to CDA. Data will be collected by community mobilisers (people affected by mycetoma trained in research methods) and overseen by the PDRFs.
4. Analysis of health promotion materials and distribution of updated materials: Health promotion materials will be collected by PDRFs from all health centres in the study site. After analysis of the materials and after relevant improvements (if necessary) are made of the materials they will be distributed at appropriate locations and events (e.g. at health centres, in community screenings, at future community engagement events). The staff and associated costs of this intervention will be analysed through a health economics questionnaire.

#### Theme 2

1. Photovoice: We will recruit 2-4 biomedical doctors, 2-4 traditional healers, and 2-4 religious healers (as relevant) to participate in photovoice from the health service providers perspective. They will photograph and document the healing journey including how they heal and treat people. Biomedical doctors will be recruited through the gatekeeper (MRC), traditional and religious healers will be identified and recruited through data collected in Theme 1. We will also recruit 10-12 participants with a new presentation or diagnosis of mycetoma to photograph and document their health process. They may be recruited from the community mobilisers or identified through stakeholder and asset mapping in Theme 1.
2. Photo-exhibition: Exhibitions will be held in public spaces with the exact location to be determined through consultation with the CAB and research participants. We aim for 150 people to attend the exhibition, albeit it might be held at more than one location. Participants will be recruited through media campaigns and word of mouth.

#### Theme 3

1. Script development: We will recruit 6-12 participants to help develop forum theatre scripts. They may be recruited from the community mobilisers or identified through stakeholder and asset mapping in Theme 1.
2. Actors and facilitator: We will recruit 6-12 actors and 1-2 facilitators to perform the scripts. Actors and facilitators will be identified through word of mouth, and from stakeholder and asset mapping in Theme 1.
3. Participants at TfD performances: Performances will be held in public spaces with the exact location to be determined through consultation with the CAB. We aim for 150 people to attend performances, albeit they might be held at more than one location. Participants will be recruited through media campaigns and word of mouth.

#### Theme 4: Health economics

Participants for the health economics questionnaire on access to health care services by people with mycetoma and associated costs will be recruited through the CDA recruitment. When a participant is asked to consent to participate in CDA we will also ask 100-150 people for consent to participate in this health economics questionnaire. Thus, the participants approached for recruitment will be the same for both the CDA and health economics questionnaire. The questionnaire will be administered by community mobilisers and analysed by the post-docs.

### Description of all processes, interventions, comparisons

The research themes will be explored using different methods, some familiar, some socio-scientific, and some creative. We aim for this methodological approach to be adaptable to other countries where mycetoma is endemic, for the anthropological study of other NTDs, and for a blueprint of intervention activities that can support early detection of mycetoma (and potentially other NTDs), thus reducing disease burden.

#### Theme 1: Understanding the disease

1. Stakeholder and asset mapping (knowledge generation);
2. Cultural Domain Analysis interviews with stakeholders on disease knowledge, attitude, and practice, and knowledge awareness (knowledge generation);
3. Identification and analysis of health promotion materials (knowledge generation), work with stakeholders to improve materials and/or develop new ones (intervention);
4. Work with local health authorities to print and distribute materials to stakeholders and at appropriate locations/events, including photovoice exhibition and TfD performances (intervention).

We foresee that the postdoctoral research fellow based in Sudan will be best equipped to carry out the strategies and methods needed to explore this theme. First, they will conduct stakeholder mapping to identify the relevant stakeholders and gatekeepers in the field site.

Stakeholders might include:

community leaders;
community organisations;
local health centres;
religious organisations and/or religious leaders;
local political leaders and regional politicians;
minority group advocates (e.g. disability, gender);
youth organisations/leaders;
Doctors, nurses, and healthcare providers.

This is only a sample of potential stakeholders, and the field research may easily identify and involve others. Specific attention will be given to the identification of representatives from the most vulnerable populations affected by mycetoma. Next, they will conduct asset mapping to identify the resources and skills of stakeholders within the field site. They might also identify physical structures or places (e.g. health centres, religious centres), businesses, associations, and institutions that can help facilitate early detection of mycetoma. The ‘assets’ will then be organised on a map within predefined community boundaries. We will ask stakeholders to participate in a workshop to co-produce a theory of change.

Second, CDA interviews with stakeholders will be conducted. In this study, CDA will explore how people living in mycetoma endemic areas think about and interpret disease contraction, preventative measures, disease risk, and who has knowledge on and can treat mycetoma. CDA is ‘the study of how people in a group think about lists of things that somehow go together. The goal is to understand how people in different cultures (or subcultures) interpret the content of domains differently’. It is a type of structured interviewing method that is very productive because it is known to be enjoyable to administer, and it is an easy way to collect and analyse data from a large number of people. Patients who attend a community-based screening at their health centre will be recruited to participate in CDA. We will recruit and train community mobilisers (people affected by mycetoma) to help conduct CDA with supervision and guidance from the postdoctoral research fellows.

Within CDA the researchers will utilise the tools of free listing, pile-sorts, and ranking. When this method is conducted in-person we will ask participants (patients and providers) to write their answers on notecards. Free listing is a simple method where participants are asked to list all they know about ‘x,’ and the goal is to get informants to list as many items as they can in a domain. We will first ask participants to list problems related to mycetoma. This question follows an exploratory approach to understanding the causes, symptoms, and consequences of mycetoma. Probing and prompting is essential in free listing and will be used carefully so as to not guide respondent answers (e.g., ‘are there any more you can think of?’). Upon completing the free listing, we will ask participants to pile sort, or to ‘put the terms together which they feel belong together’. This is a simple and compelling method for collecting data about ‘what goes with what’. We will ask participants to put their answers from the free listing together according to what they feel belongs together. The piles might include ‘how the disease is contracted,’ ‘treatments,’ ‘symptoms,’ etc. The final part of our cultural domain analysis is to ask participants to rank order their lists. We will have participants rank in order the most important problem to the least. Their answers will illuminate priorities, values, and potentially even perceived risk of the disease. We will analyse the data according to the variables of knowledge, attitude, practice, and knowledge awareness. The ranking and pile-sorting exercises will help us understand the problems people face related to mycetoma. Care will be taken to analyse not only what was listed, but also what was left off. Findings will form a base for developing the intervention(s). Data on knowledge awareness will serve as another layer to the stakeholder and asset mapping exercises. It will also guide us as to whom and where to potentially distribute improved health promotion materials.

We intend for the CDA exercise to be framed around problems and for asset mapping to highlight positives in the community. We hope to be able to make connections between the problems identified and the assets available to address them. This will allow us to make community and district level policy recommendations based on evidence.

Third, health promotion materials will be collected from relevant authorities, including health centres. We will ask for relevant mycetoma health materials from health workers with the intention to understand the perceptions of the disease by those that produce the materials and those that consume them through interviews with patients (consumers) and doctors/MOH (producers). We would like to analyse them according to the variables of intended audience, stigma, message. We will apply health promotion theories while working with stakeholders to improve them and hopefully distribute in locations identified in asset mapping, in ‘knowledge awareness,’ as well as at photovoice workshops and exhibitions, and to TfD audiences. The methods in this theme will be structured and conducted in the above order to allow for an iterative approach where tools and questions can be refined based on knowledge gained (and also replicable as needed for future data collection/analysis).

#### Theme 2: Traditional healers and biomedicine

1. Conduct a rapid ethnography (34,35) and interviews to understand the role of religious healing in health seeking behaviours (knowledge generation);
2. Carry out photovoice with traditional healers, biomedical doctors, and patients in three distinct groups. Modify the groups to potentially include religious healers based on the findings of step one (knowledge generation and intervention);
3. Upon the conclusion of photovoice, bring the three groups together for a wider discussion about health seeking behaviour and early detection of mycetoma. Encourage and support/facilitate actionable (and sustainable) recommendations between the groups (knowledge generation and intervention);
4. Hold a photovoice exhibition in an appropriate public location(s) to engage the public in conversation about early detection (intervention)

The Sudan-based PhD student will conduct their research on this topic and lead the photovoice intervention as part of their project. They will begin by conducting a rapid ethnography and interviews with stakeholders to better understand religious healing. They will draw on the literature on rapid research in disease outbreaks to inform their approach: they will (a) analyse usage of religious healers in regards to mycetoma and health needs in general, (b) evaluate mycetoma control strategies through religious healing tools/methods and compare to other methods like traditional healing, (c) identify infrastructure and resources used in religious healing including specific prayers and mass prayer gatherings, and (d) identify of the causes of mycetoma and transmission from a religious perspective.

Next, they will carry out photovoice with three separate groups: traditional healers, doctors, and patients. Then, these methods and others will be further utilised to enable these individuals to come together for group discussion and the co-creation of intervention strategy and ways forward.

Finally, they will hold a photo exhibition in locations deemed appropriate by stakeholders and in line with our asset mapping. The aim is to facilitate public conversations about mycetoma in an effort to reduce stigma, improve health promotion, improve knowledge of mycetoma, and inform the public on appropriate knowledge awareness.

#### Theme 3: Stigma

1. Employ narrative research approaches to collect stories of felt, enacted, and structural stigma from patients and their family members (knowledge generation and intervention);
2. Identify and train TfD actors in drama techniques as well as community mobilisation (intervention);
3. Develop forum and playback theatre script and performance (knowledge generation and intervention);
4. Work with project collaborators to develop a film and/or radio drama (intervention)

First, narrative research approaches will be conducted to collect stories of felt, enacted, and structural stigma from patients and their family members. We have chosen the narrative approach as there is evidence that narratives are effective in reducing stigma and narratives constructed with the first-person point of view are ‘superior’ to reducing stigma (36). Stigma reduction is vital in encouraging early presentation of mycetoma. Additionally, narratives are ‘possessed with a capacity for social justice, which allows historically marginalized and silenced peoples to tell their stories and for others to listen and respond’ (37). Thus, a narrative approach lends itself to being a precursor to TfD.

Narrative research is the exploration of stories to make sense of lived experience. It entails the intimate engagement of the research in the story of the relationship, in other words, the researcher is written into the work. This approach demands reflexivity and authentic self-reflection. It employs interview techniques over a long period of time that consider temporality (a past, present, and future), sociality (relationship between researcher and participant), and place (a sequence of locations or one location).

Next, the doctoral student will facilitate TfD to allow individual and community stories to be told to a wider audience. TfD is a tool used to improve the quality of life among vulnerable and marginalised populations. It uses fiction and the ‘safe space’ of performance to comment on reality and offer alternatives. There are two types of theatre in TfD: forum and playback. Forum theatre is based on Boal’s Theatre ‘Theatre of the Oppressed’ and is an interactive theatre form in which audience members identify their ‘internal oppressions’ in order to begin to overcome them. The audience is shown a play in which the main character encounters an obstacle which s/he is unable to overcome. The subject matter, in this case stigma, is based on a shared life experience. When the play has been performed members of the audience can suggest alternative options for how the protagonist could have acted. The obstacle can be rehearsed to uncover alternatives to the situation. The actors explore the results of these choices with the audience creating a debate in which experiences and ideas are shared, generating solidarity and a sense of empowerment. Playback theatre is when someone in the audience tells a moment or story from their life then chooses the actors to play the different roles. Next, everyone present at the ‘theatre’ watches the enactment as the story “comes to life” with artistic shape and nuance. Actors can draw on many styles to convey meaning, including metaphor or song.

To conduct TfD the doctoral student must first themselves be trained in the method and either support the training of or train themselves the actors. We recommend Prof. Mufunangi Magalasi from the University of Malawi to carry out the training as he is in our network of collaborators. The doctoral student will identify the appropriate people to take part and the best locations to hold the theatre. The actors will be identified based on their motivation to not only act in the performances but to also serve as community mobilisers. We hope the actors will include patients with or recovered from mycetoma, perhaps even patients from the Mycetoma Cultural Village, and that acting and serving as community mobilisers will be income generating activities. The data from the scripts and the performances will be analysed to understand how theatre can reduce stigma and encourage early presentation. Finally, the TfD scripts will be developed into a film and radio drama, both of which can live beyond the life of the study.

#### Theme 4: Health economics

1. We will collect the data on contacts with health professionals (doctors, nurses, health assistants, pharmacists), the use of laboratory services, and hospitalisations. We will also collect information about the cost of medication. We will ask participants about contacts with traditional and religious healers and expenses associated with these contacts, including travel, food, accommodation and costs for the accompanying person(s). The data will be collected using bespoke questionnaires based on our recent study (Phase 1). Data will be collected during the first phase of data collection (‘understanding the disease’) and participants will be recruited through the CDA recruitment. The questionnaire will be administered by community mobilisers and analysed by the post-docs.
2. We will measure the generic HR-QoL in people with mycetoma using the World Health Organization tool WHOQOL-BREF, Arabic version (38). WHOQOL-BREF asks respondents how they feel about their quality of life and health, including physical, psychological, social, and environmental health. It has five response levels (e.g. “not at all”, “slightly”, “moderately”, “very” and “extremely”) and yields a multi-dimensional profile of scores across domains and sub-domains of quality of life. WHOQOL is one of the most rigorously evaluated instruments to provide valid scores for comparison across different settings (39). We will also collect WHOQOL scores for the general population to compare these to people affected by mycetoma). Data will be collected during the third phase of data collection (‘stigma’) and asked of participants attending the TfD performances. Data will be collected by community mobilisers and analysed by the post-docs.

### Outcomes

The primary effectiveness outcome will hopefully be improved mycetoma presentation and diagnosis across all sectors of healers. This outcome will be measured by the corresponding and parallel research conducted by other work packages in the NIHR Research Unit on NTDs. The following secondary effectiveness outcomes include:

1. Enhanced knowledge and awareness
2. Reduced stigma
3. Better health-related quality of life
4. Improved health-seeking behaviour

We also intend for the following outputs to be developed to support the outcomes:

1. **Website** to showcase our qualitative study, the stages of the intervention, and to act as a home for our visual/auditory outputs
2. **TfD scripts** which can be reimagined through future performances or different creative mediums
3. **Radio drama** based on TfD scripts to be circulated in mycetoma endemic areas and to potentially be played at treatment centres, like the MRC, when patients are waiting to be seen by a doctor
4. **Film** based on TfD scripts.
5. **Photographs** from photovoice for public exhibitions in both Sudan and U
6. **Health promotion materials** to be distributed in locations determined in asset mapping
7. **Publications**:
8. Intervention implementation and CICI process evaluation as a means to encourage early presentation
9. TfD as a potential method to reduce mycetoma stigma and to be used for other NTDs
10. Photovoice as a tool to encourage early presentation and to be used for other NTDs
11. Findings from CDA
12. **Policy recommendations** for early presentation and stigma reduction

### Data management plans

Data protection will be ensured throughout the study process, from data collection to data storage, analysis and publication. Structured, multi-level precautions will be taken to safeguard the confidential nature of the information collected, and to ensure the anonymity of participants. Data collectors will be fully trained to understand and implement ethical and research governance safeguards and protections; they will undergo GDPR training, as well as in-depth training in proper data management, including maintenance of confidentiality and ensuring privacy during data collection. Data will be utilised in conformity with the written and oral consent provided by participants. All data will be collected in Sudan. Only pseudonymised data will be shared between the teams in BSMS and Sudan. With the permission of participants, interviews will be tape-recorded and assigned personal identifiers (PIDs); interviews will then be transcribed in Sudanese Arabic and translated into English. If a translator is needed, they will be hired from outside the community, to ensure anonymity of participants. Tapes will be kept securely within the unit they originated from and wiped clean after completion of the study. Electronic versions of pseudonymised transcripts will be kept securely. Names and contact details of participants will be collected in written consent forms and will be kept securely; they will not be transferred to the UK. PIDs will be used; identifying data will be kept in a separate list with identifying codes used on the research data. Any person-identifiable data will not leave the unit from which they originated, and keys to identification numbers will be held confidentially and securely within the respective research unit, separately from the research data collected. The document linking participant identification numbers with personal data will only be accessible to the study research team in Sudan. Only pseudonymised data will be used for data analyses.

### Safety considerations

Safety considerations include the possibility that the nature of the research could potentially have an emotionally disturbing impact on the researcher as researchers will hear about negative experiences and adverse medical outcomes related to mycetoma. To address this the community mobilisers will have access to support with the MRC in both Sinnar State and in Khartoum. They will also have the opportunity to discuss their feelings, worries, etc. in regular group discussions with the PDRFs. The PhD student has access to support through BSMS, their supervisors, and the MRC. The PDRFs also have access support through BSMS, senior academics in the NIHR Unit, and the MRC. Additionally, researchers may conduct the research alone in an official office (e.g. local ministry of health, NGO). In response researchers will have a database of all research activities, times, and locations that the research team has access to. Researchers will inform their supervisors of upcoming research activities (time, date, location) and ensure the entire research team has their phone number.

### Analysis

#### Theme 1

1. Stakeholder mapping: stakeholders will be analysed according to

a. List of actors and influencers in mycetoma treatment, policy, activism, etc.
b. Physical proximity to lead organisations and treatment centres
c. Links and relationships between stakeholders
d. Stakeholder influence and attitude (ranked)
e. Prioritising top stakeholders Results will be analysed by the PDRFs and the PhD student.
2. Asset mapping: community assets will be analysed according to

a. The assets of individuals: these are their skills, knowledge, networks, time, interests and passions
b. The assets of associations: this is not just the formal community organisations or voluntary groups. It includes all the informal networks and ways that people come together
c. The assets of organisations: this is not just the services that organisations deliver locally, but also the other assets they control
d. The physical assets of the study site including buildings, transport, etc.
e. The economic assets of the study site including how stakeholders are contributing to the local economy and how this can influence early diagnosis of mycetoma
f. The cultural assets of the study site including mapping for talents and opportunities for everyone to express themselves Results will be analysed by the PDRFs and the PhD student.
3. CDA: CDA data will be analysed according to the variables of knowledge, attitude, practice, and knowledge awareness. Care will be taken to analyse not only what was listed, but also what was left off. Data on knowledge awareness will serve as another layer to the stakeholder and asset mapping exercises. It will also guide us as to whom and where to potentially distribute improved health promotion materials. Data will be analysed by PDRFs.
4. Health promotion materials: Health promotion materials will be analysed according to the variables of

a. Medical: is the content scientifically accurate?
b. Behavioural change: does the content influence behaviour change (positive/negative)? Was this long-lasting or temporary?
c. Educational: is the content educational and appropriate for the intended audience?
d. Client centred: who is the intended audience and are the materials appropriate for them? Did the materials reach the community it was intended to reach?
e. Societal change: is the community more aware of the message now than they were before?
f. Stigma: is the content stigmatising and how can we make it inclusive? Data will be analysed by PDRFs on Theme 4C.
5. Health economics: health economics questionnaires will be analysed by the PDRFs under the supervision of (lead health economist on the NIHR Research Unit on NTDs). The analysis will be conducted in Microsoft Excel. The analysed data will include:

a. Completed patient health-related quality of life questionnaires WHOQOL-BREF (https://www.who.int/tools/whoqol) captured using REDCap (a secure web application for building and managing online surveys and databases https://www.project-redcap.org/);
b. Completed Patient Healthcare Resource Use questionnaires purposively designed and captured using REDCap;
c. Study co-design questionnaires completed by the research staff to capture resource use and costs associated with developing the intervention (Word document). The output of data analysis will include:

a. Resources and costs associated with developing and delivering the intervention.
b. WHOQOL-BREF scores for physical health, psychological well-being, social relationships and environment;
c. A number of contacts with health professionals (doctors, nurses, health assistants, pharmacists); the use of laboratory services, hospitalisations, and associated expenses (including travel, food, accommodation and costs for the accompanying person(s); a number of contacts with traditional and religious healers and associated costs; cost of medication and traditional remedies; money borrowing over the past year;

#### Theme 2

1. Photovoice: data from photovoice group discussions will be analysed according to the variables of
  a. Health seeking behaviour
  b. Health seeking pathway
  c. Confluence and divergence
2. Photography: the photographic images will be analysed (40) according to the study variables of
  a. Health seeking behaviour
  b. Health seeking pathway
  c. Confluence and divergence
  while allowing additional themes to emerge.
3. Photo-exhibition: discussions had at the photo-exhibition will be analysed according to the variables of
  a. Health seeking behaviour
  b. Health seeking pathway
  c. Confluence and divergence

#### Theme 3

1. TfD scripts and performance discussion with audience will be analysed according to the variables of

a. Stigma: perceived and experienced stigma; factors that enable stigma perpetuation
b. Demographics: Gender, age, occupation

Ethical considerations:

This study’s ethical review at BSMS RGEC has been paused until the situation in Sudan becomes more stable and it is safe to carry out global health research.

### Timeline

This research will be conducted over a series of four stages.

#### Stage 1: Establishing Phase 2 of the NIHR Global Health Research Unit on NTDs

In this stage we developed a study design and protocol with partners from the MRC. We gained gatekeeper approval to ensure the success of the study. We have recruited a postdoctoral research fellow and PhD student, both based in Sudan. Finally, we have obtained ethical approval from the Research Governance and Ethics Committee at Brighton and Sussex Medical School.

#### Stage 2: Development of the study themes and thematically organised data collection

In this stage we will begin initial data collection for theme one by identifying stakeholders, conducting asset mapping, and establishing a community advisory board who will help develop a theory of change. In theme two we will identify participants for photovoice and provide them initial training in basic camera use. In theme three we will identify TfD actors and training will be provided to the actors and relevant staff. We will begin to develop the scripts and work on the practicalities of where and how to carry out the TfD. Finally, we will hold a check-in for all research team members to identify challenges, discuss solutions, assess progress, and determine appropriate next steps.

#### Stage 3: Extension and further development of data collection; problem-solving; completion of data collection

In this stage we will complete all data collection. In theme we will collect and analyse health promotion materials. Then we will redistribute updated materials to health centres and at engagement events, namely photovoice exhibitions and theatre performances. In theme two we will conduct photovoice and in theme three we will carry out the TfD performances. Again, we will conduct a check-in to assess our progress, problem solve, and begin discussing data analysis themes that are emerging across all themes.

#### Stage 4: Analysis, writing-up and publication; next steps

In this stage we will focus on analysing the data by theme and across the entire medical anthropology component of the study. We will write up the data by theme and wholistically resulting in a study publication and several theme-based publications. We will produce reports for funders and potentially a brief for stakeholders. We will also discuss the future extension and expansion of research.

## Discussion

Through the research and community engagement activities described above, this study aims to support the early presentation and diagnosis of mycetoma in Sinnar, Sudan. This will be achieved by engaging stakeholders and key community assets, by developing a Community Advisory Board to provide guidance and feedback throughout the research process as well as to help develop a theory of change, and by engaging those directly affected by mycetoma through the role of community mobiliser. We will also implement several community engagement interventions to reduce stigma, provide increased knowledge of mycetoma, and to enable stakeholders to be producers of knowledge. This will be achieved through improvement of health promotion materials, a photo-exhibition, and theatre performances.

This study faces several limitations including potentially not being able to reach the most vulnerable and marginalized. There is a risk this study will primarily include participants who are mobile and able to visit public locations when many people suffering with mycetoma might be homebound due to age, chronic health conditions, or amputation related to the disease. We will only be able to include those willing to disclose their experience with mycetoma, a severely stigmatized disease. There may be some people in the community who are currently hiding their diagnosis to protect their social and economic standing. There is also the risk of researcher bias, however we hope to overcome this through an international team of five academic researchers working on this medical anthropology component alone. We also hope the CAB will provide guidance and reflect any bias back to us as researchers.

The health economics component will help us to understand what healthcare services are used by people with mycetoma. This information will support decision-makers in allocating scarce healthcare resources. We will also pilot the questionnaires for measuring health-related quality of life (the main effectiveness outcome in economic evaluations). These data will inform our future cost-effectiveness studies of interventions to improve early detection and diagnostics of mycetoma.

The findings from this study will be disseminated through academic publications, briefs shared with stakeholders, and reports and presentations shared with the entire NIHR Global Health Research Unit. Community engagement interventions also act as a dissemination mechanism for findings generated by participants to be presented back to the community in creative and inclusive ways, namely photography and theatre.

Amendments to the study will firstly be submitted to the BSMS RGEC. Upon ethical approval the protocol will be updated, and the research timeline modified.

## Data Availability

N/A

## Authors’ contributions

CA- conceptualization, methodology, writing-original draft and preparation, writing-review and editing

VH- conceptualization, methodology, writing-original draft and preparation

ES- writing-review and editing

NH- conceptualization and methodology of the health economics component, writing-original draft and preparation

ME- writing-review and editing

SZ- conceptualization, methodology, writing-review and editing, supervision

SB- writing-review and editing

## Acknowledgements

Thank you to Gail Davey and Melanie Newport for your insightful and constructive comments.

## Footnotes

1 For example, for those most in need, visiting the MRC in Khartoum can take a whole day or days due to distance from their homes. As well as travel being costly, these distances are limiting to the extent that individuals do not feel that it is possible to go there for treatment - at least not regularly, as is required for effective treatment of mycetoma infection.

